# Protocol for the DREAMER study: design and methodological framework of a multicenter trial-ready cohort of individuals with isolated REM sleep without atonia

**DOI:** 10.64898/2026.05.15.26353348

**Authors:** Raffaele Ferri, Monica Puligheddu, Michela Figorilli, Giuseppe Plazzi, Fabio Pizza, Luigi Ferini-Strambi, Sara Marelli, Giuseppe Lanza

## Abstract

Isolated rapid eye movement sleep behavior disorder is a strong clinical marker of future alpha-synucleinopathy, but earlier stages of this risk pathway remain insufficiently characterized. Rapid eye movement sleep without atonia is the polysomnographic substrate of this disorder and may also be detected in individuals without clinical dream-enactment behavior. Whether isolated rapid eye movement sleep without atonia is a benign finding or an early risk state for future rapid eye movement sleep behavior disorder and neurodegeneration remains unknown. DREAMER is a multicenter, prospective, observational cohort protocol designed to identify adults without clinical rapid eye movement sleep behavior disorder who show isolated rapid eye movement sleep without atonia during full-night laboratory video-polysomnography. Four Italian sleep centers will use harmonized eligibility criteria, standardized clinical and sleep assessment, quantitative REM Atonia Index scoring, secure web-based data capture, and planned longitudinal follow-up. Adults aged 40 years or older undergoing video-polysomnography will be screened. Participants with prior rapid eye movement sleep behavior disorder or technically inadequate REM sleep/chin electromyographic data will be excluded. Isolated rapid eye movement sleep without atonia will be defined in participants without clinical rapid eye movement sleep behavior disorder using a REM Atonia Index threshold of <0.85. The target recruitment is more than 500 participants over 18 months, with an expected enriched subgroup of approximately 85 individuals with isolated rapid eye movement sleep without atonia. Ancillary neurophysiological assessments and blood sampling for future biomarker studies will be obtained when feasible. DREAMER is intended to create a harmonized, trial-ready cohort for evaluating isolated rapid eye movement sleep without atonia as a potential early risk marker for incident rapid eye movement sleep behavior disorder and subsequent neurodegenerative outcomes. The study is registered at ClinicalTrials.gov as DREAMER, ClinicalTrials.gov Identifier NCT06140511.

## 1. Introduction

REM sleep behavior disorder (RBD) is characterized by dream-enactment behavior associated with loss of physiological muscle atonia during REM sleep [1]. When RBD occurs in the absence of an established neurological disorder or other clearly causative condition, it is commonly regarded as an isolated form of RBD and has become one of the most powerful clinical markers of prodromal alpha-synucleinopathy. Longitudinal studies have shown that a large proportion of patients with isolated RBD eventually develop Parkinson disease, dementia with Lewy bodies, or multiple system atrophy, often after a prolonged interval [1-3].

The central diagnostic polysomnographic feature of RBD is REM sleep without atonia (RSWA). However, RSWA is not limited to clinically manifest RBD [4]. Quantitative PSG studies have shown that isolated RSWA/RWA can be detected in individuals without clinical RBD or dream-enactment behavior, with age, sex, medication exposure, psychiatric status, sleep-disordered breathing, and referral context influencing its frequency and interpretation [5-9]. This raises an important but unresolved question: does isolated RSWA represent a benign polysomnographic variant, a medication-related phenomenon, or an early risk state along the pathway toward RBD and synucleinopathy? Small longitudinal and retrospective sleep-clinic studies have suggested that isolated RSWA may be associated with subsequent RBD or neurodegenerative outcomes, whereas more recent registry-linked data emphasize that iRSWA is heterogeneous and not equivalent to iRBD [10-12]

This question has direct implications for clinical trial design. Neuroprotective and disease-modifying trials in alpha-synucleinopathies increasingly require identification of individuals at very early stages, ideally before overt motor or cognitive neurodegenerative syndromes emerge. Isolated RBD already provides a high-risk population, but it may not represent the earliest detectable stage. If isolated RSWA identifies at least a subgroup of individuals upstream of RBD, it could widen the prevention window and provide a biomarker-enriched population for future risk-stratified trials [4, 10, 11]. At present, however, isolated RSWA lacks standardized prospective cohort infrastructure, harmonized multicenter ascertainment, and longitudinal evidence sufficient to support its use as a trial-enrichment criterion.

The DREAMER study, Isolated REM sleep without Atonia as a risk factor for REM sleep behavior disorder, is designed to address this methodological gap. DREAMER is not an interventional clinical trial. Rather, it is a multicenter, prospective, registry-like cohort created to identify individuals with isolated RSWA among adults undergoing laboratory video-polysomnography (vPSG), harmonize their clinical and neurophysiological characterization, and establish a long-term follow-up platform to determine whether isolated RSWA predicts incident RBD and, eventually, alpha-synucleinopathy. The study is explicitly designed as trial-enabling infrastructure: it standardizes eligibility definitions, quantitative RSWA assessment, electronic data capture, biomarker collection, and risk-management procedures required for future preventive or disease-modifying studies.

This article describes the rationale, design, operational methodology, data-management framework, planned analyses, and risk-mitigation strategy of DREAMER, with emphasis on elements relevant to clinical trial readiness.

## 2. Materials and Methods

### 2.1 Objectives

The primary objective of DREAMER is to establish a multicenter, harmonized, trial-ready cohort of adults without clinical RBD in whom isolated RSWA can be identified quantitatively by vPSG and followed longitudinally for the development of RBD.

The secondary objectives are:

1. To estimate the frequency of isolated RSWA in adults aged ≥40 years undergoing laboratory vPSG across participating sleep centers.
2. To compare clinical, sleep, pharmacological, and neurophysiological characteristics between participants with and without isolated RSWA.
3. To evaluate whether quantitative RSWA severity, measured by the REM Atonia Index (RAI), is associated with clinical and instrumental markers potentially relevant to prodromal synucleinopathy.
4. To create a secure, quality-controlled electronic dataset suitable for future long-term phenoconversion analyses.
5. To establish a biobank and multimodal biomarker framework that can support future mechanistic and prevention-oriented studies.

The long-term objective, beyond the 24-month cohort-construction period, is to determine whether baseline isolated RSWA is associated with higher risk of incident RBD and subsequent neurodegenerative outcomes compared with absence of RSWA.

### 2.2. Study design

DREAMER is a multicenter, prospective, observational cohort with registry-like data architecture. The study includes a 24-month phase dedicated to protocol harmonization, ethics approval, electronic case report form (eCRF) implementation, recruitment, baseline vPSG-based classification, and collection of ancillary biomarker data. Long-term follow-up is planned after completion of recruitment. The overall DREAMER workflow, from screening to longitudinal outcome ascertainment and trial-ready cohort construction, is shown in Fig 1, and the core design elements of DREAMER and their relevance to future trial readiness are summarized in Table 1.

**Table 1.**
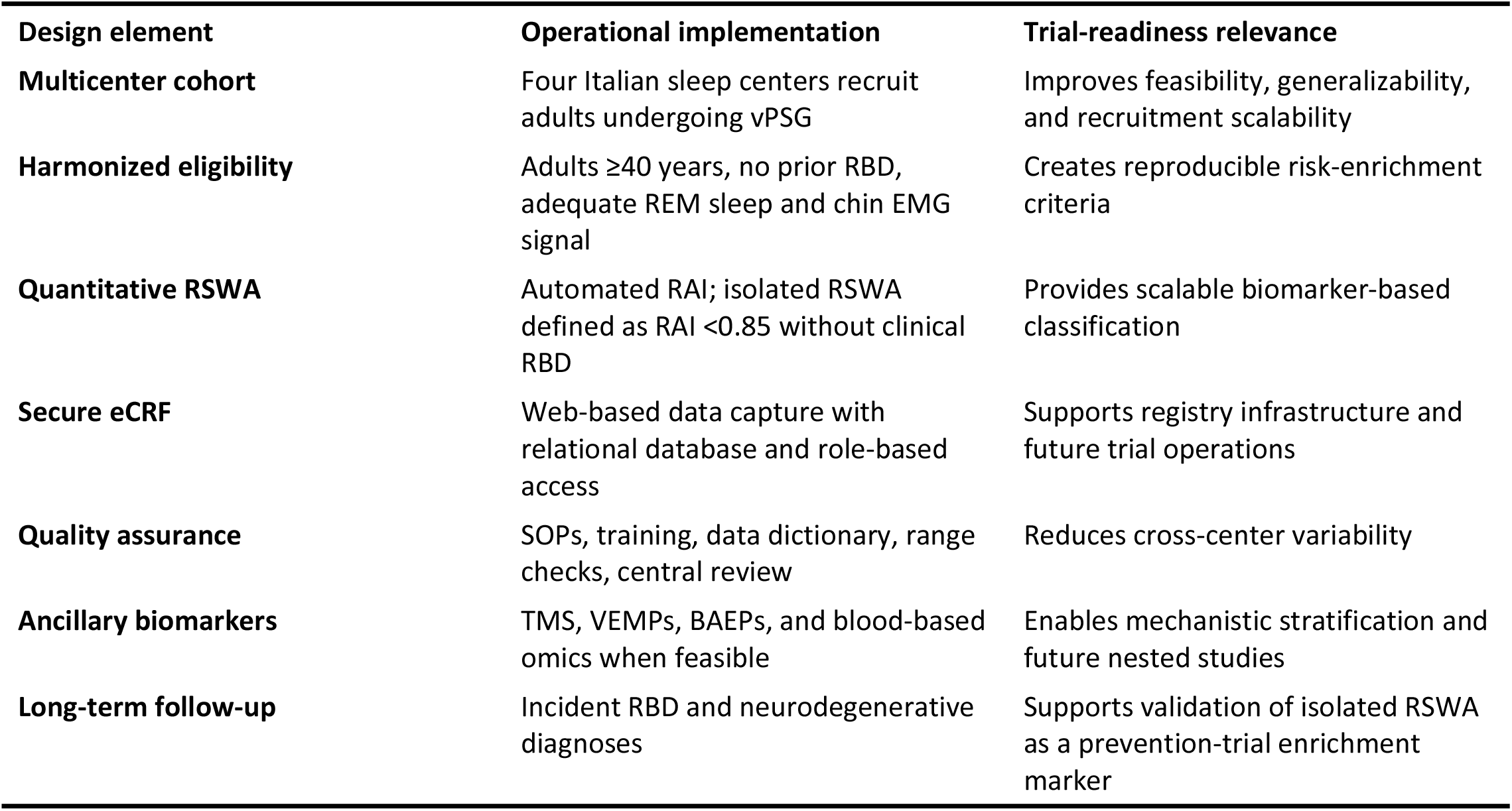
DREAMER design elements.

**Fig 1.**
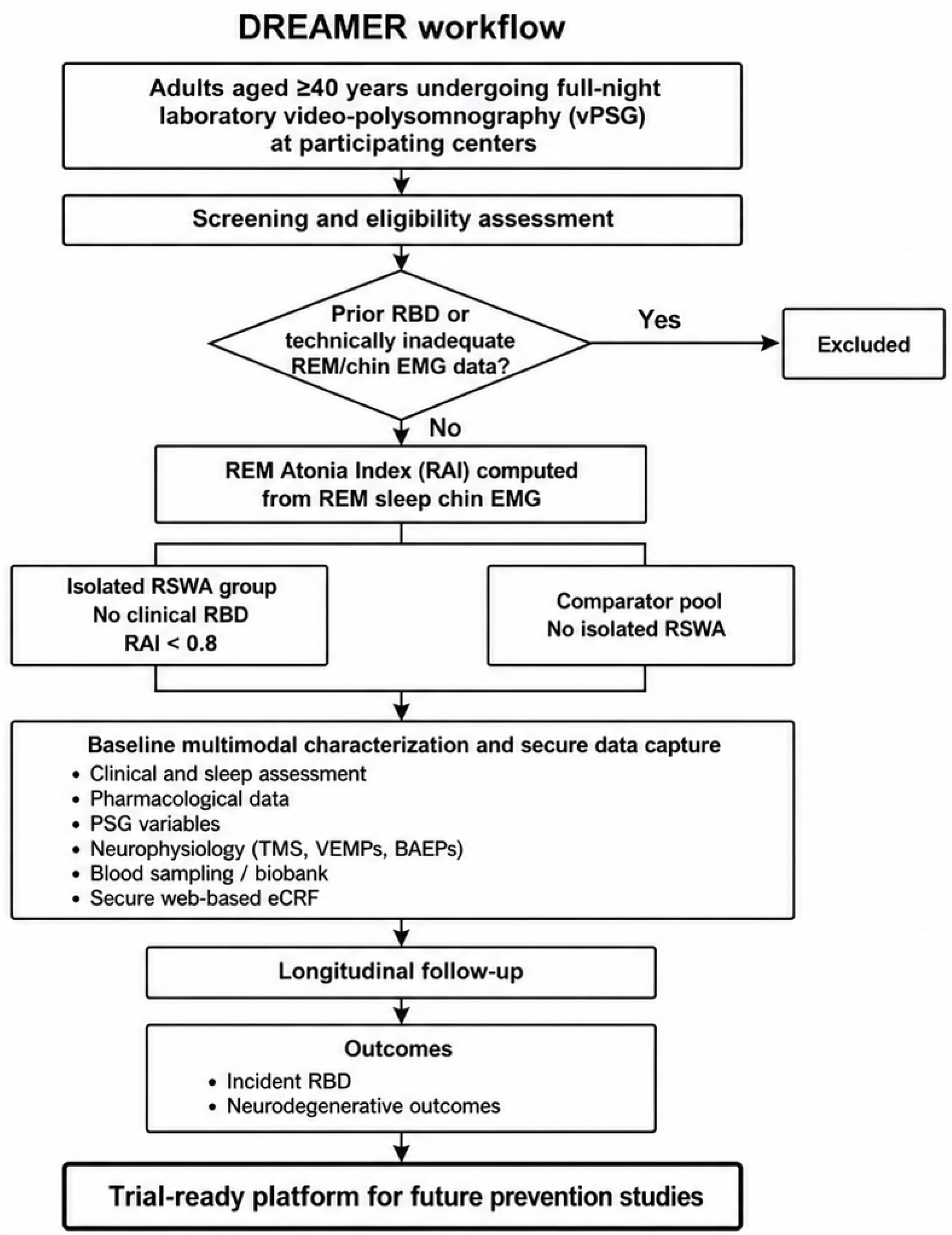
DREAMER study workflow. Adults aged ≥40 years undergoing full-night laboratory video-polysomnography are screened for eligibility. After exclusion of participants with prior RBD or technically inadequate REM/chin EMG data, the REM Atonia Index is computed to classify participants with isolated RSWA and comparator participants without isolated RSWA. Baseline multimodal characterization, secure eCRF-based data capture, and longitudinal follow-up support the creation of a trial-ready platform for future prevention studies.

The study has three operational phases:

**Phase 1: Harmonization and infrastructure development**. During the initial months, participating centers will agree on eligibility criteria, clinical variables, vPSG procedures, RAI computation, ancillary neurophysiological protocols, blood-sample collection procedures, data dictionary, and quality-control rules. A secure web-based eCRF will be implemented.

**Phase 2: Recruitment and baseline phenotyping**. Adults undergoing full-night laboratory vPSG will be screened. Participants without prior RBD and with technically adequate REM sleep recordings will be classified according to quantitative RAI values. Clinical, pharmacological, sleep, neurophysiological, and biobanking data will be collected according to the shared protocol.

**Phase 3: Long-term follow-up**. Participants with isolated RSWA and a comparator group without RSWA will be followed to identify incident RBD and neurodegenerative outcomes. The follow-up component is intended to continue beyond the initial funding period and to provide the evidentiary basis for future risk-stratified prevention trials.

The schedule of screening, baseline, ancillary, and follow-up assessments is shown in Table 2.

**Table 2.** Schedule of assessments.

| Study phase | Procedure | All enrolled participants | Isolated RSWA subgroup |
| --- | --- | --- | --- |
| Screening | Eligibility review and informed consent | X | X |
| Baseline | Clinical and sleep history | X | X |
| Baseline | Medication and comorbidity assessment | X | X |
| Baseline | Full-night laboratory vPSG | X | X |
| Baseline | RAI computation | X | X |
| Baseline | RBD symptom/questionnaire assessment | X | X |
| Baseline | TMS | As feasible | Priority |
| Baseline | VEMPs/startle response | As feasible | Priority |
| Baseline | BAEPs | As feasible | Priority |
| Baseline | Blood sampling for biobank | As feasible | Priority |
| Follow-up | Annual assessment for dream-enactment behavior | Planned | X |
| Follow-up | Repeat vPSG | If clinically indicated or protocol-defined | X |
| Follow-up | Neurodegenerative outcome ascertainment | Planned | X |

### 2.3. Setting and participating centers

Four Italian sleep centers participate in DREAMER:

1. Oasi Research Institute-IRCCS, Troina, Italy, serving as coordinating and recruiting center.
2. University Hospital of Cagliari, Sleep Disorders Research Center, serving as recruiting center and neurophysiology site.
3. IRCCS Istituto delle Scienze Neurologiche di Bologna, serving as recruiting center and brainstem neurophysiology site.
4. IRCCS Ospedale San Raffaele Turro, Milan, serving as recruiting center and brainstem neurophysiology site.

All participating centers have established expertise in sleep medicine, RBD assessment, vPSG acquisition and scoring, and clinical neurophysiology. The multicenter structure is intended to increase recruitment feasibility, improve generalizability across sleep-clinic populations, and enable harmonized data collection across distinct operational environments.

### 2.4. Study population, inclusion and exclusion criteria

Participants will be recruited among adults undergoing full-night laboratory vPSG for clinical indications at participating centers. The target population is intentionally pragmatic: it reflects real-world sleep-center populations in which incidental RSWA may be detected and future risk stratification may become clinically relevant.

Participants must meet all of the following criteria:

1. Age ≥40 years at baseline.
2. Ability to understand study procedures and provide written informed consent.
3. Undergoing full-night laboratory vPSG with video recording.
4. Availability of chin electromyographic signal of sufficient technical quality for RAI computation.
5. At least 5 minutes of artifact-free REM sleep.
6. No prior clinical diagnosis of RBD.

Collateral history from a bed partner, roommate, or other reliable informant will be collected whenever available to improve ascertainment of dream-enactment behavior. This is particularly important because the distinction between iRSWA and iRBD may be influenced by the availability of an observer or co-sleeper, potentially leading to under-recognition of dream-enactment behavior [12].

Participants will be excluded if any of the following are present:

1. Prior diagnosis of RBD.
2. Severe neurological, psychiatric, cognitive, or general medical condition that prevents valid participation or understanding of study procedures.
3. vPSG recordings with major artifacts affecting the chin EMG signal during REM sleep.
4. Less than 5 minutes of artifact-free REM sleep.
5. Withdrawal of consent.

The protocol will record medications known to influence REM sleep muscle tone, particularly antidepressants, as well as comorbid sleep disorders and neurological conditions. These factors will not automatically exclude participation unless specified by the final protocol or judged to preclude valid classification, but they will be incorporated into sensitivity analyses.

### 2.5. Baseline assessment

At baseline, all participants will undergo standardized clinical assessment, including demographic characteristics, medical history, neurological history, psychiatric history, cognitive status when clinically indicated, sleep history, history of dream-enactment behavior, sleep-related injuries, parasomnia symptoms, and medication exposure. Particular attention will be paid to antidepressants, dopaminergic medications, benzodiazepines, melatonin, antipsychotics, and other drugs that may influence REM sleep physiology or motor behavior during sleep.

Sleep history will include symptoms of insomnia, excessive daytime sleepiness, obstructive sleep apnea, restless legs syndrome, periodic limb movements, narcolepsy-related symptoms when relevant, and other parasomnias. Standardized RBD screening questionnaires may be used as supportive tools but will not replace clinical and vPSG-based classification.

All participants will undergo full-night laboratory vPSG according to the procedures routinely used in participating centers and harmonized through the DREAMER protocol. Recordings will include electroencephalography, electrooculography, submental electromyography, electrocardiography, respiratory channels, limb electromyography, oxygen saturation, body position, and synchronized video. Sleep staging and respiratory and movement events will be scored according to the current standards adopted by each center and harmonized for the variables included in the DREAMER eCRF.

The key classification variable is RSWA quantified by the RAI. The RAI is an automated measure derived from submental EMG amplitude during REM sleep and has been proposed as part of a practical approach to improve diagnostic accuracy and accessibility in RBD assessment [13]. Values range from 0, indicating complete absence of EMG atonia, to 1, indicating stable EMG atonia [14, 15]. The DREAMER protocol adopts the established three-level RAI classification: >0.9 normal, 0.8-0.9 mildly abnormal, and <0.8 definitely abnormal [14-16]. Participants with RAI <0.85 and no clinical RBD will be classified as having isolated RSWA, while sensitivity analyses will explore alternative RAI thresholds, including <0.8.

The RAI algorithm will be applied blind to clinical classification whenever feasible. Because the RAI is computed automatically, no manual adjustment of the algorithmic output will be permitted. Technical adequacy checks will focus on chin EMG quality, artifact burden, and the minimum REM sleep duration required for valid computation.

### 2.6. Ancillary neurophysiological and biomarker assessments

DREAMER incorporates ancillary assessments to generate mechanistic and trial-readiness information beyond vPSG [17].

The coordinating center will perform transcranial magnetic stimulation (TMS) in participants with isolated RSWA and in an appropriate comparison group without RSWA. TMS will assess cortical excitability and inhibitory/facilitatory intracortical mechanisms. The aim is to determine whether isolated RSWA is associated with early cortical neurophysiological changes analogous to those described in RBD or early synucleinopathy [18].

The Cagliari center will perform cervical, masseter, and ocular vestibular-evoked myogenic potentials when feasible. These measures provide information on brainstem pathways that may be relevant to REM atonia regulation and early RBD-related neurophysiology [19]. Startle response assessment may also be included according to the final harmonized protocol.

The Bologna and San Raffaele centers will perform brainstem auditory evoked potentials in selected participants with isolated RSWA and matched comparators without RSWA. These measures will complement other neurophysiological assessments of brainstem function.

Blood samples will be obtained whenever possible, with priority given to participants with isolated RSWA. Samples will be processed and stored for future proteomic, metabolomic, and glycomic analyses. The purpose is to determine whether isolated RSWA is associated with peripheral molecular signatures overlapping with those previously described in isolated RBD [20, 21].

Brain magnetic resonance imaging, computed tomography, dopamine transporter imaging, or other investigations will not be mandated by the protocol, but may be performed when clinically indicated as part of routine diagnostic work-up. Results will be recorded in the eCRF when available.

### 2.7. Outcomes

The main outcomes of the 24-month cohort-construction phase are:

1. Number of participants screened, eligible, enrolled, and classifiable by RAI.
2. Frequency of isolated RSWA among enrolled participants.
3. Completeness and quality of vPSG, clinical, neurophysiological, and biosample data.
4. Feasibility of harmonized multicenter eCRF-based data capture.
5. Recruitment rate and retention readiness for long-term follow-up.

The primary long-term clinical outcome is incident RBD, defined by development of clinical dream-enactment behavior with vPSG evidence of RSWA according to accepted diagnostic criteria.

Secondary long-term outcomes include:

1. Change in RAI over time, when repeat vPSG is available.
2. Development of subthreshold dream-enactment behavior or increased RBD questionnaire scores.
3. Diagnosis of Parkinson disease, dementia with Lewy bodies, multiple system atrophy, or other neurodegenerative disorders.
4. Association between baseline RAI and neurophysiological markers.
5. Association between baseline RAI and blood-based proteomic/metabolomic/glycomic signatures.
6. Attrition, loss to follow-up, and feasibility of repeated annual assessment.

The procedures used to identify and confirm these outcomes during follow-up are described below.

### 2.8. Follow-up procedures and phenoconversion ascertainment

Participants classified as having isolated RSWA and a comparator group without isolated RSWA will be invited to participate in longitudinal follow-up. Follow-up will be performed at least annually by in-person visit, telephone interview, video consultation, or review of clinical records, according to participant availability and local center procedures. The follow-up strategy is designed to detect two clinically relevant forms of phenoconversion: conversion to clinically manifest RBD and conversion to a neurodegenerative disorder within the alpha-synucleinopathy spectrum, in line with longitudinal evidence from isolated RBD cohorts and biomarker-based models of conversion risk [2, 3].

At each follow-up contact, participants will be asked about the occurrence of new dream-enactment behavior, vocalizations or complex motor behaviors during sleep, sleep-related injuries, falls from bed, violent or disruptive sleep behaviors, and changes in sleep quality. Whenever possible, information will also be obtained from a bed partner, caregiver, roommate, or other reliable informant. Follow-up assessments will also record changes in medication exposure, especially antidepressants, benzodiazepines, melatonin, dopaminergic medications, antipsychotics, and other drugs potentially influencing REM sleep muscle tone or parasomnia expression. New diagnoses of sleep disorders, neurological disorders, psychiatric disorders, and relevant systemic illnesses will be recorded.

Incident RBD will be suspected when new recurrent dream-enactment behavior or other clinically relevant REM sleep-related motor behaviors are reported by the participant or an informant. In these cases, participants will be invited to undergo repeat full-night laboratory video-polysomnography, including chin electromyography and video documentation, whenever feasible. Incident RBD will be defined by the emergence of clinical dream-enactment behavior together with polysomnographic evidence of RSWA, according to accepted diagnostic criteria [1, 22]. For each suspected or confirmed case, the date of first reported symptoms, date of clinical recognition, and date of polysomnographic confirmation will be recorded separately.

Phenoconversion to neurodegenerative disease will be assessed through annual clinical history, neurological review, and documentation of any new diagnosis of Parkinson disease, dementia with Lewy bodies, multiple system atrophy, or other relevant neurodegenerative disorders, using established diagnostic frameworks where applicable [23-25]. Follow-up data collection will include symptoms or signs suggestive of parkinsonism, cognitive decline, visual hallucinations, fluctuating cognition, autonomic dysfunction, gait impairment, olfactory impairment, and other non-motor manifestations potentially relevant to prodromal synucleinopathy, consistent with research criteria and updates for prodromal Parkinson disease [26, 27]. This approach is also supported by longitudinal application of prodromal Parkinson disease criteria in RBD cohorts [28].

When clinically indicated, participants will be referred for neurological evaluation and additional investigations according to local clinical practice, including neuropsychological testing, autonomic assessment, brain imaging, dopamine transporter imaging, or other biomarker investigations. These tests are not mandated by the protocol but will be recorded when available.

All suspected phenoconversion events will be reviewed by the recruiting center and, when necessary, discussed centrally by the study investigators. Outcome adjudication will use predefined criteria and will distinguish between no conversion, possible RBD, confirmed RBD, possible neurodegenerative phenoconversion, and confirmed neurodegenerative diagnosis. For longitudinal analyses, the event date will be defined as the earliest reliable date of symptom onset when available; otherwise, the date of clinical diagnosis or polysomnographic confirmation will be used. Participants who cannot be contacted will be classified as lost to follow-up after repeated contact attempts, and the date of last available assessment will be recorded.

### 2.9. Sample size and recruitment assumptions

DREAMER uses a pragmatic recruitment target based on expected RSWA frequency and feasibility within participating centers. Previous quantitative work in adults without RBD showed that isolated RSWA elevations may be detected in a substantial minority of individuals, with approximately 14% of patients without dream-enactment behavior reaching previously established RBD-level RSWA thresholds [4, 7]. Therefore, the initial recruitment target is >500 participants over 18 months, expected to yield approximately 85 individuals with isolated RSWA. Because the DREAMER population is restricted to adults aged ≥40 years, and because RSWA becomes more frequent with age, the number of isolated RSWA cases may exceed this estimate.

The initial 24-month phase is not designed as a definitive phenoconversion trial. Instead, it is designed to assemble an enriched cohort and collect the standardized baseline data required for adequately powered follow-up. Recruitment assumptions will be reassessed after the first 6 months of enrollment. If the observed frequency of isolated RSWA or the rate of technically classifiable vPSG recordings is lower than expected, recruitment targets may be increased, and the comparator strategy may be adjusted by selecting an age- and sex-matched subgroup without RSWA for long-term follow-up.

Assuming approximately 85 participants with isolated RSWA and long-term follow-up over 5 years, the study may observe a sufficient number of incident RBD events to generate preliminary estimates of relative risk compared with participants without RSWA. These estimates will be used to plan future prevention or neuroprotection trials rather than to support immediate clinical implementation.

### 2.10. Data management and quality assurance

A web-based eCRF will be developed by the coordinating center. The data architecture will include a relational database for storage, a secure web service for data management, and a client-side web application for authorized users. The system will support data entry, display, modification, and controlled deletion according to predefined user permissions.

Data will be pseudonymized before entry into the central database. Access will be role-based and password-protected. The system will be hosted on secure institutional infrastructure. Privacy, cybersecurity, and data-protection procedures will be supervised with involvement of data-protection and information-technology expertise at the coordinating institution.

A data dictionary will define all mandatory, optional, and derived variables. Harmonized fields will include demographic data, vPSG variables, RAI values, sleep-disorder diagnoses, RBD-related symptoms, neurological history, medication exposure, neurophysiological test results, biosample status, and follow-up outcomes.

Quality-control procedures will include:

1. Standard operating procedures for vPSG acquisition and RAI computation.
2. Training meetings among centers before recruitment.
3. Predefined rules for technical adequacy of chin EMG and REM sleep duration.
4. Automated eCRF range checks and missing-data flags.
5. Periodic cross-center data-quality reports.
6. Central review of ambiguous classification cases.
7. Documentation of protocol deviations.

### 2.11. Statistical analysis plan

Baseline characteristics will be summarized for the overall cohort and stratified by RSWA status. Continuous variables will be reported as mean and standard deviation or median and interquartile range, according to distribution. Categorical variables will be reported as counts and percentages.

Participants with isolated RSWA will be compared with participants without RSWA. Continuous variables will be analyzed using Student’s t test or Mann-Whitney U test, as appropriate. Categorical variables will be analyzed using chi-square or Fisher’s exact tests. Effect sizes and confidence intervals will be reported whenever possible to avoid overreliance on significance testing.

RAI will be analyzed both as a continuous variable and as a categorical classification variable. Multiple linear regression will evaluate associations between RAI and clinical, pharmacological, sleep, and neurophysiological predictors. Logistic regression will evaluate predictors of isolated RSWA status. Candidate covariates will include age, sex, antidepressant exposure, neurological comorbidity, sleep apnea severity, REM sleep duration, and participating center.

For long-term follow-up, cumulative incidence of RBD will be described using Kaplan-Meier methods. Cox proportional hazards models will estimate the association between baseline isolated RSWA and incident RBD, adjusted for age, sex, medication exposure, center, and relevant sleep-disorder comorbidities. Competing-risk methods may be considered for incident neurodegenerative outcomes or death. Sensitivity analyses will evaluate the influence of antidepressant exposure, minimum REM duration, comorbid sleep apnea, and alternative RAI thresholds.

The extent and pattern of missing data will be reported. If missingness is limited, complete-case analyses will be used. If missingness is substantial and plausibly missing at random, multiple imputation may be considered for regression models. Follow-up attrition will be summarized and compared between RSWA and non-RSWA groups.

### 2.12. Study status and timeline

The DREAMER study was registered at ClinicalTrials.gov before completion of recruitment. Ethics approval was obtained on 24 January 2024. Participant recruitment started on 22 February 2024 and completed on 18 May 2026. Longitudinal follow-up and outcome data collection are ongoing and are planned to continue for 5– 10 years after baseline assessment. The first analyses of baseline cohort characteristics and isolated rapid eye movement sleep without atonia frequency are expected by 30 August 2026, whereas longitudinal analyses of incident rapid eye movement sleep behavior disorder and neurodegenerative outcomes are expected after completion of the planned follow-up period, approximately by 2031.

### 2.13. Ethical and regulatory considerations

DREAMER will be conducted in accordance with the Declaration of Helsinki, applicable national regulations, and institutional ethics requirements. The study protocol was approved by the Comitato Etico Locale IRCCS Oasi Maria SS on 24 January 2024 (approval number CEL-IRCCS OASI/24-01-2024/EM02). Written informed consent will be obtained from all participants before inclusion. Because the study is observational and non-interventional, risks are primarily related to confidentiality, routine video-polysomnography burden, ancillary neurophysiological procedures, and blood sampling. Participants will receive standard clinical management for any clinically relevant findings emerging from routine care. The study was registered at ClinicalTrials.gov as DREAMER – IsolateD REM Sleep Without Atonia as a Risk Factor for REM Sleep Behavior disordER (ClinicalTrials.gov Identifier: NCT06140511; Unique Protocol ID: PNRR MR1 2022-12375694).

## 3. Discussion

DREAMER addresses a critical methodological barrier in the development of prevention trials for synucleinopathies: the need to identify and characterize individuals at risk before overt RBD or neurodegenerative disease develops. Isolated RBD is already a strong marker of future alpha-synucleinopathy, but it may be too late in the prodromal sequence for some preventive strategies. Isolated RSWA is biologically plausible as an earlier stage, because RSWA is the PSG substrate of RBD and may appear before clinically evident dream enactment; however, available longitudinal data remain limited and partly divergent [10-12]. However, isolated RSWA cannot yet be used as a trial-enrichment marker without prospective evidence, harmonized measurement, and reproducible cohort infrastructure.

The design of DREAMER is therefore deliberately trial-oriented, despite its observational nature. It incorporates several features relevant to future clinical trials: a prespecified eligibility framework, standardized vPSG-based classification, quantitative automated RSWA measurement, centralized eCRF architecture, predefined data-quality checks, multicenter recruitment, collection of confounders, and ancillary biomarker assessments. These elements will facilitate later conversion of the cohort into a risk-enriched platform for observational follow-up, nested case-control analyses, and potentially future preventive or disease-modifying trials.

The use of RAI is a central methodological feature. Manual RSWA scoring is informative but may be time-consuming and variable across laboratories. A quantitative automated index provides a scalable approach that can be applied across centers [14-16, 29] and may support reproducible risk classification. The selected threshold of RAI <0.85 is intended to enrich the cohort for abnormal REM atonia loss while retaining sufficient sensitivity for a prospective risk-identification study; sensitivity analyses using more stringent thresholds, including RAI <0.8, will be important.

DREAMER also incorporates multimodal neurophysiology. TMS may identify cortical excitability changes related to inhibitory and facilitatory circuits. VEMPs and BAEPs may capture brainstem dysfunction relevant to REM atonia regulation. Blood-based omics may reveal peripheral signatures overlapping with those previously reported in isolated RBD. Although these ancillary assessments are exploratory, they may help determine whether isolated RSWA is merely a polysomnographic finding or part of a broader prodromal neurobiological profile.

Several limitations should be acknowledged. First, DREAMER recruits from sleep-center populations rather than the general population; therefore, RSWA frequency estimates may not be population prevalence estimates. Second, isolated RSWA may be heterogeneous, reflecting age, sex, antidepressant or psychoactive medication exposure, psychiatric status, sleep fragmentation, respiratory events, neurological comorbidities, observer availability, or subclinical RBD [5, 7-9, 12]. Because sleep-disordered breathing may complicate RSWA interpretation, sensitivity analyses stratified by sleep apnea severity and respiratory-event-related arousals will be important [30]. The protocol addresses this through systematic collection of confounders and sensitivity analyses, but residual confounding is likely. Third, the 24-month funded phase is primarily a cohort-construction period and cannot by itself provide definitive phenoconversion evidence. Fourth, long-term follow-up requires sustained funding, participant retention, and harmonized outcome adjudication. Finally, RBD diagnosis during follow-up may depend on repeat vPSG availability, which may vary across centers and over time.

Despite these limitations, DREAMER is positioned to make a methodological contribution. Contemporary trial development in neurodegeneration increasingly depends on earlier risk detection, biomarker enrichment, scalable data infrastructure, and harmonized multicenter operations. DREAMER provides a framework for moving isolated RSWA from an incidental PSG observation toward a systematically characterized risk state that can be evaluated for future clinical trial enrichment.

## 4. Conclusions

DREAMER is a multicenter, prospective, registry-like cohort designed to identify isolated RSWA in adults without RBD using harmonized vPSG and quantitative RAI assessment. By integrating standardized clinical assessment, secure eCRF-based data capture, neurophysiological testing, biobanking, and planned long-term follow-up, DREAMER aims to build trial-ready infrastructure for evaluating isolated RSWA as an early risk marker in the RBD-synucleinopathy continuum. If isolated RSWA proves to predict incident RBD, this framework may support earlier enrollment windows for future preventive and neuroprotective trials.

## Data availability

No datasets were generated/analyzed for this article and de-identified study data will be made available after completion, subject to ethics/privacy restrictions, data-protection regulations, and study-governance agreements.

## Author contributions

R.F. conceived the study and coordinates the DREAMER project. R.F., G.L., M.P., G.P., and L.F.S. contributed to study design and multicenter organization. G.L., M.P., M.F., F.P., and S.M. contributed to the neurophysiological, PSG, and biomarker components. R.F. and G.L. drafted the manuscript. All authors critically revised the manuscript for intellectual content and approved the final version.

## Declaration of generative AI and AI-assisted technologies in the manuscript preparation process

During the preparation of this work, the authors used ChatGPT, OpenAI, to improve readability and language. After using this tool, the authors reviewed and edited the content as needed and take full responsibility for the content of the manuscript.

## Acknowledgments

The authors thank the participating centers and study personnel involved in the implementation of the DREAMER protocol.

